# Prevalence and Recurrence of Pica Behaviors in Early Childhood: Findings from the ALSPAC Birth Cohort

**DOI:** 10.1101/2023.06.04.23290951

**Authors:** Natalie M. Papini, Cynthia M. Bulik, Samuel JRA Chawner, Nadia Micali

## Abstract

**Objective:** Pica has been largely understudied in general population samples. Pica occurs most often in childhood and appears more prevalent in individuals with autism and developmental delays (DD). Pica occurrence in the general population is poorly understood due to few epidemiological studies.

**Method:** Data on 10,109 caregivers from the Avon Longitudinal Study of Parents and Children (ALSPAC) study who reported pica behavior at 36, 54, 66, 77, and 115 months on their child were included. Autism was obtained through clinical and education records, while DD was derived from the Denver Developmental Screening Test.

**Results:** A total of 312 parents reported pica behaviors in their child. Of these, 19.55% reported pica at least at two waves (n=61). Pica was most common at 36 months (N=226; 2.29%) and decreased as children aged. A significant association was found between pica and autism at all five waves (p < .001). There was a significant relationship between pica and DD, with individuals with DD more likely to experience pica than those without DD at 36 (p = .01), and 54 (p < .001), 65 (p=.04), 77 (p <.001), and 115 months (p=.006). Exploratory analyses examined pica behaviors with broader eating difficulties and child body mass index.

**Discussion:** Pica is an uncommon behavior in childhood; however, children with DD or autism may benefit from pica screening and diagnosis between ages 36-115 months. Children who exhibit undereating, overeating, and food fussiness may also engage in pica behaviors.

---

Pica is a feeding and eating disorder in DSM-5, and is defined as recurrent intake of non-nutritive, non-food substances (e.g., paper, soap) for at least one month, is inappropriate to the developmental stage of the individual, occurs outside of cultural norms, and occurs in the absence of another mental or physical condition (American Psychiatric Association, 2013). Pica is relatively understudied; however, individuals who experience pica frequently have adverse health and psychosocial outcomes (APA, 2013). Individuals with pica are at increased risk of significant medical consequences linked to dental enamel erosion, infection, anemia, and gastrointestinal obstruction from ingested non-food substances (APA, 2013). In addition to significant medical risks, pica is associated with diminished psychosocial well-being, specifically a lack of strong and supportive relationships with family, and less social contact (Ashworth et al., 2009).

Pica can occur at any point in life, with onset most commonly during childhood (APA, 2013). Common co-occurrences with pica include autism and developmental delays (DD), including intellectual disabilities (ID) (APA, 2013; Fields et al., 2021). Pica behavior, which involves the consumption of non-nutritive foods, differs from a formal pica diagnosis, which is based on the diagnostic criteria outlined in the DSM-V. Fields and colleagues (2021) assessed pica in children 30 to 68 months of age and found 3.5% of population-based controls reported pica behavior. Roughly 0.3-25% of children with IDs demonstrated pica behavior, while 14-36% of children with autism have shown pica behavior (Ali, 2001; Fields et al., 2021; LoVullo & Matson, 2009). Pica behaviors have been reported in 28% of children with autism and ID (Fields et al., 2021). Additionally, children from lower socioeconomic backgrounds are more likely to develop pica (Leung & Hon, 2019).

Notable gaps in pica research exist. First, the extent of pica occurrence within community settings is poorly understood because there are few epidemiological studies (Sturmey & Williams, 2016). Of the few studies examining pica prevalence using general population data, none have examined longitudinal data to investigate how prevalence changes across different developmental stages and persists across development. Our goal was to address this research gap by utilizing prospective data gathered from a population cohort to study pica behaviors over time from a sample representative of the general population.

Research from the Avon Longitudinal Study of Parents and Children (ALSPAC) has yielded significant findings related to child and adolescent eating behaviors and eating disorders and, therefore, can provide essential insights into pica. Previous work using data from ALSPAC examined feeding, diet, and growth of children with autism and found differences in feeding symptoms at infancy and a less varied diet (with fewer vegetables, salad, fruit, sweets, and carbonated beverages) at 15 months compared with controls (Emond et al., 2010). This work also reported that children with autism were more likely than controls to report pica behaviors at 38 and 54 months. To date, no studies have examined the stability of pica behaviors throughout childhood using longitudinal prospectively reported data. Other ALSPAC research has assessed eating disorder diagnosis across three time points throughout adolescence but did not include pica (Micali et al., 2017). Furthermore associations between childhood eating behaviors during the first ten years of life and eating disorder diagnoses and behaviors at age 16 were observed using ALSPAC self-report and parent-reported data (Herle et al., 2020). However, this work primarily focused on the development of binge-eating behaviors, binge-eating disorder, and anorexia nervosa (Herle et al., 2020), and did not include pica.

The present study reports the prevalence, longitudinal stability, and correlates of pica in children in the ALSPAC cohort. We evaluated pica behaviors across five waves, including questionnaires completed by mothers at 36, 54, 65, 77, and 115 months. Additionally, we compared the prevalence of pica between individuals with and without autism and DD. An exploratory aim of the present study was to determine the co-occurrence of pica and child eating behaviors, including food fussiness (the tendency to only consume specific foods with refusal to try new foods; Dovey et al., 2008), overeating, and undereating. Furthermore, we examined if pica behavior is related to child body mass index (BMI).

## Methods

### Ethical approval

This study involves secondary analysis of data initially collected in the Avon Longitudinal Study of Parents and Children (ALSPAC) study. Informed consent for the use of data collected via questionnaires and clinics was obtained from participants following the recommendations of the ALSPAC Ethics and Law Committee at the time. Ethical approval for the study was obtained from the ALSPAC Ethics and Law Committee and the Local Research Ethics Committees.

### Participants

All pregnant women living in Avon, UK who were expected to deliver between April 1, 1991, and December 31, 1992, were eligible to participate in the Avon Longitudinal Study of Parents and Children (ALSPAC) study (Boyd et al., 2013; Fraser et al., 2013). ALSPAC is a longitudinal, population-based prospective study of pregnant women and their children. All women provided informed and written consent. The study website contains details of all the vailable data through a fully searchable data dictionary and variable search tool. To access, reference the following webpage: http://www.bristol.ac.uk/alspac/researchers/our-data/.

Analysis of the ALSPAC sample was limited to participants in which child sex at birth was reported and where pica data was available in at least two of five waves (n=10,109) to examine the persistence of pica behaviors over time (defined as pica-reported behaviors at two or more waves).

### Measures

#### Pica

At 36, 54, 65, 77, and 115 months, mothers were asked, “How often does the child eat coal, dirt, or other non-nutritious substances?” Response options included “every day,” “at least once a week,” “less than once a week,” and “not at all.” Pica behavior in the current study was defined as any respondent option that indicated pica occurred “every day,” “at least once a week,” and “less than once a week.” When examining differences in pica between subgroups, a binary “yes/no” pica variable was created.

#### Early Childhood Developmental Skills

Child development was measured at 30 months using an adapted version of the Denver Developmental Screening Test (DDST-II) (Frankenburg & Dodds, 1967). The DDST-II has been used in ALSPAC to examine early child development (Hameed et al., 2018). The DDST-II examines four categories of development: social skills, fine motor skills, communication, and gross motor skills. Questions were adapted to be appropriate to the child’s age and were completed by the mother. The 30-month assessment was selected because it preceded the first pica assessment. For analyses in the current study, the total development score at 30 months was recoded into a new variable: those in the 10^th^ percentile (indicating DD) and those outside of the 10^th^ percentile (indicating no DD). Other researchers have used ALSPAC developmental measures to create a binary developmental problems measure using the 10^th^ percentile as a marker for DD (Riglin et al., 2017).

#### Autism

Autism diagnosis derived from a previous ALSPAC study was accessed (Williams et al., 2008). This previous study derived autism diagnosis from clinical and educational records (Williams et al., 2008), and ethical approval for this was obtained from the ALSPAC Law and Ethics committee and local research ethics committees (NHS Haydock REC: 10/H1010/70. At age 18, study children were sent “fair processing” materials describing ALSPAC’s intended use of their health and administrative records. They were given clear means to consent or object via a written form. Data were not extracted for participants who objected or were not sent fair processing materials.

#### Eating Behaviors

Mothers answered items regarding child eating behaviors: food fussiness, overeating, and undereating. Food fussiness was measured at each of the five waves (36, 54, 65, 77, and 115 months) included in this study. Three items measured food fussiness: 1) “How worried are you because your child is choosy,” 2) “How worried are you because your child has feeding difficulties,” and 3) “How worried are you because your child is refusing food.” Respondents were instructed to answer these items with the following options: “no/did not happen,” “not worried,” “a bit worried,” and “greatly worried.” Overeating was measured using a single item: “How worried are you that your child is overeating?” Similarly, undereating was captured with a single item: “How worried are you because your child is undereating?” Both items contained the same answer options: “yes, very worried,” “yes, a bit worried,” “yes, not worried,” and “no, didn’t happen.” Dichotomous variables were created for food fussiness, overeating, and undereating at all five waves. Individuals who selected any of the following answer options: “yes, very worried,” “yes, bit worried,” “yes, not worried,” were coded as 1. Those who selected “no, didn’t happen” were coded as 0. A total food fussiness variable was derived from selecting any confirmed food fussiness (i.e., any response of “yes, very worried,” “yes, a bit worried,” “yes, not worried”) on any of the three food fussiness variables.

#### Child Body Mass Index (BMI) Z-Scores

Child BMI data were collected at 37, 49, 61, 102, and 120 months. BMI z-scores were used to examine the relationship between BMI and pica. To determine if pica behaviors predict child BMI, a linear regression was conducted for months 36, 54, and 66. Because of fewer pica cases in months 77 and 115, comparisons were made using independent samples t-tests to examine if there were significant differences in BMI z-scores between children who reported pica behaviors and those who had not reported pica behaviors.

#### Data Analysis

First, we analyzed the prevalence of pica across the five waves where pica was available to better understand pica prevalence over time. Next, we compared differences in the presence of pica behaviors between participants based on sex (male/female), those with and without autism, and those with and without a DD (defined as below the 10^th^ percentile of the total development score at 30 months). Chi-square or Fisher’s exact tests were conducted for categorical data; Fisher’s exact test was used when any expected cell count was less than 5. Alpha values were set at .05. Because these analyses were exploratory, no adjustment was made for multiple comparison testing. This should be considered when interpreting findings.

## Results

### Pica Prevalence Across Five Waves

Pica frequency was the highest at 36 months (N=226; 2.29%) and decreased in prevalence across childhood, including 54 (0.78%), 65 (0.62%), 77 (0.55%), and 115 months (0.33%). (see Table 1).

**Table 1.** Pica Prevalence Across Five Data Collection Waves

|  | Pica Waves |  |  |  |  |
| --- | --- | --- | --- | --- | --- |
|  | 36 Months<br>n/N (%; 95 CI) | 54 Months<br>n/N (%; 95 CI) | 65 Months<br>n/N (%; 95 CI) | 77 Months<br>n/N (%; 95 CI) | 115 Months<br>n/N (%; 95 CI) |
| Overall prevalence | 226/9,881<br>(2.29%;<br>2.0%-2.6%) | 74/9,497<br>(0.78%;<br>0.6-1.0%) | 54/8,758<br>(0.62%;<br>0.5-0.8%) | 46/8,361<br>(0.55%;<br>0.4-0.7%) | 25/7,650<br>(0.33%;<br>0.2-0.5%) |
| Within Male<br>Sex at Birth | 119/5,097<br>(2.33%) | 38/4,909<br>(0.77%) | 29/4,515<br>(0.64%) | 25/4,296<br>(0.58%) | 19/3,875<br>(0.49%) |
| Within Female<br>Sex at Birth | 107/4,784<br>(2.24%) | 37/4,588<br>(0.81%) | 25/4,243<br>(0.59%) | 21/4,065<br>(0.52%) | 6/3,775<br>(0.16%) |
| Autism Present | 8/64 (12.5%) | 7/63 (11.11%) | 6/59 (10.17%) | 8/59 (13.6%) | 6/56 (10.71%) |
| Autism not<br>Present | 218/9802<br>(2.22%) | 66/9,418<br>(0.70%) | 48/8,684<br>(0.55%) | 38/8,290<br>(0.46%) | 19/7,584<br>(0.25%) |
| DD Present | 28/794<br>(3.53%) | 14/775<br>(1.81%) | 9/773<br>(1.16%) | 11/695<br>(1.60%) | 6/615<br>(0.98%) |
| DD not Present | 150/7,198<br>(2.08%) | 39/6,980<br>(0.55%) | 27/6,510<br>(0.41%) | 24/6,254<br>(0.38%) | 12/5,662<br>(0.21%) |

*Note.* n=cases, N=total sample, CI= Confidence Interval, DD= developmental delay

### Pica Associations with Sex at Birth

Chi-square goodness of fit analyses were used to determine the relationship between pica and sex at birth (female/male). There was a significant relationship between pica and child sex at birth at 115 months; the prevalence of pica was greater in males than females (0.49% vs. 0.18%), *X*^2^ (1, *N* = 6,330) = 6.01, *p* = .01). No associations in pica behaviors were observed between male or female sex at 36, 54, 65, or 77 months.

### Pica Associations with Autism

Fisher’s exact test was applied to determine if pica presence was associated with autism diagnosis. A significant association was found between pica and autism at all five waves with pica being more prevalent in autistic children (p < .001) (see Table 1).

### Pica Associations with DD

Both chi-square goodness of fit and Fisher’s exact test were used to examine the relationship between pica and DD at each of the five waves. Fisher’s exact test was used at months 65, 77, and 115 when the expected cell count violated the assumption of 5 or greater needed for chi-square. There was a significant relationship between pica and DD, with individuals who had DD more likely to experience pica than those without DD at 36 months, *X*^2^ (1, *N* = 7,992) = 6.83, *p* = .01, and 54 months, *X*^2^ (1, *N* = 7,755) = 16.00, *p* < .001. Fisher’s exact test indicated significant relationships between pica and DD at 65 (p=.04), 77 (p <.001), and 115 months (p=.006).

### Longitudinal Stability of Pica

A total of 312 parents reported pica behaviors in their child. Of these, 19.55% reported pica at least at two waves (n=61) (Table 2). Pica onset was most common at 36 months (N=226; 2.29%) and decreased in prevalence across childhood (See Table 1). A total of 24 participants (0.24%) reported pica at 115 months (Time 5). Nine individuals reported pica at 36 months and again at 115 months but did not report pica behaviors at 54, 65, or 77 months. This indicates that pica behaviors persisted over 6.5 years (from 36 months to 115 months) and fluctuated (did not report pica at all 5 waves) for 9 out of 61 individuals (14.75%).

**Table 2.** Longitudinal Stability of Pica

*Longitudinal Stability of Pica*
|  | Number of assessment waves pica persisted |  |  |  |
| --- | --- | --- | --- | --- |
|  | 1 wave | 2 waves | 3 waves | 4 or more waves* |
| Frequencies | 251 | 41 | 12 | 8 |

*Note.* This table presents the number of assessment waves an individual reported pica behavior.
\*In accordance with ALSPAC's requirements not to allow cell counts <5, "4 waves" and "5 waves" categories were combined into "4 or more waves".

### Co-occurring Features

#### Association of Pica with other eating behaviors and BMI

Logistic regression models were conducted to examine the relationship between food fussiness, overeating, and undereating with pica behaviors at 36, 54, and 65 months. These models controlled for child sex at birth. Of thesebehaviours, undereating and overeating were significantly associated with pica at 36 months. All three behaviours (food fussiness, overeating, and undereating) were significantly associated with pica at 54 months. None of the three eating behaviors were significantly associated with pica at 65 months. Because pica cases decreased over time, we were not sufficiently powered to run logistic regression at later waves (77 and 115 months). As such, chi-square goodness of fit or Fisher’s tests were used at 77 and 115 months to examine the relationship between these eating behaviors and pica. Chi-square goodness of fit tests indicated overeating, undereating, and food fussiness were not significantly related to pica at 77 months. Both overeating *X*^2^ (1, *N* = 5,824) = 4.24, *p* = .04, and undereating *p* < .001 were significantly associated with pica at 115 months (see Table 3).

**Table 3.** Associations between sex, child eating behaviors, and pica at 36, 54, 65, 77, and 115 months

| | Odds Ratio (95% CI) | | | $X^2$ | |
| --- | --- | --- | --- | --- | --- |
|  | 36 months | 54 months | 65 months | 77 months | 115 months |
| *Sex*<br>*(female)* | 0.93 (0.71-1.22)<br>*p*=0.74 | 1.02 (0.64-1.63)<br>*p*=0.95 | 0.96 (0.56-1.65)<br>*p*=0.75 | 0.16<br>*p*=0.70 | 6.45<sup>+</sup><br>*p*=0.01 |
| **Child Eating Behaviors** |  |  |  |  |  |
| Food Fussiness | 1.14 (0.76-1.70)<br>*p*=0.53 | 0.45 (0.25-0.83)<sup>+</sup><br>*p*=-0.01 | 1.62 (0.72-3.64)<br>*p*=0.25 | 2.47<br>*p*=0.12 | 0.50<br>*p*=0.5 |
| Undereating | 2.04 (1.50-2.78)<sup>+</sup><br>*p* < .001 | 2.07 (1.24-3.47)<sup>+</sup><br>*p*=0.01 | 1.21 (0.63-2.34)<br>*p*=0.57 | 0.00<br>*p*=0.99 | 12.74<sup>+</sup><br>*p* < .001 |
| Overeating | 1.48 (1.08-2.04)<sup>+</sup><br>*p*=0.01 | 1.74 (1.02-2.96)*<br>*p*=0.04 | 1.65 (0.86-3.15)<br>*p*=0.13 | 1.60<br>*p*=0.21 | 4.24*<br>*p*=0.04 |

*Note.* CI= Confidence Interval, \*= significant at the $\leq .05$ level, += significant at the $\leq .01$ level.

#### Child BMI and Pica

Results from linear regression models at 36, 54, and 65 months indicated no statistically significant association between pica and child BMI. Similarly, independent samples t-tests indicated no significant difference in BMI between children with and without pica at 77 (pica BMI z-score =.1389 (n=35), no pica BMI z-score = .1242 (n=6,378), p= 0.47) and 115 months (pica BMI z-score =-.1659 (n=11), no pica BMI z-score = -.0015 (n=3,827), p= 0.22).

## Discussion

In the ALSPAC cohort pica was reported by mothers in 2.3% of their children at 36 months and the behavior became less common as children aged. Sex differences in pica prevalence were not present early in development but a significant difference emerged at 115 months when pica behaviors were reported to be more common in males. We observed significant associations of pica with autism and with DD at all waves of data collection. Of children who experienced pica at any time point, 20% had recurrent pica (i.e., pica was reported in at least one other time point). Lastly, we examined the relationship between pica prevalence and child BMI and child eating behaviors (food fussiness and undereating/overeating). Pica was found to be associated with increased likelihood of broader eating difficulties such as undereating, overeating, and food fussiness. No associations were observed between the presence of pica and child BMI.

The prevalence of pica within the ALSPAC cohort is lower than that reported in adults (5.5% of adults; Hartmann et al., 2022), but broadly similar to that reported in other samples of children (3.5%; in the population-based control group; Fields et al., 2021). The higher percentage of adults endorsing pica behaviors reported by Hartmann and colleagues (2022) may be explained by differences in pica measurement. In the present study, pica frequencies are reported by the child’s caregiver, who observed pica behavior over the five time points assessed. In contrast, Hartmann and colleagues used a 7-point Likert scale where 0 indicated “never true,” and six indicated “always true.” Pica behavior was defined as any respondent who endorsed 1-6 on this Likert scale. Pica measurement used in Hartmann and colleagues (2022) may have led to higher pica reports since scores of 1-3 reflected subthreshold pica behaviours (where 0 is no pica and scores of 4-7 indicate pica).

The present study observed no evidence for a consistent significant association between sex at birth and pica behaviors at the majority of waves. Only at 115 months (age 9) were males more likely than females to exhibit pica, and it should be noted that the prevalence difference was small (age 9, 0.49% in males vs. 0.16% in females). These findings do not align with those reported by Fields and colleagues (2021), who examined children in children aged 7-14 years old with autism, other developmental disabilities, and general population-based controls and found significant differences in pica between males and females ages 48-68 months.

Further, significantly more caregivers of male children than female children ages 7-14 years old reported pica prevalence, and caregivers of male children reported higher frequencies of pica (selection of “often or greater” in response to “I like to eat things that are not meant for eating” item on the Eating Disorders in Youth Questionnaire (EDY-Q); Hilbert & van Dyck, 2016). The present study also observed significantly more males than females engaging in pica behaviors at age 9. Longitudinal research is needed to understand when sex differences emerge amongst children ages 3-14 years old using population-level data. Additional research that covers a wider range of ages could facilitate a better understanding of who is most at-risk and the optimal time to intervene.

Autism was associated with pica behaviors across all waves in this study. This association is aligned with other ALSPAC findings, which showed children with autism were more likely than controls to demonstrate pica behavior at 38 and 54 months (Emond et al., 2010). Emond and colleagues (2010) focused on children with autism and examined ALSPAC time points at 6, 15, 24, 38, and 54 months. The current study expands upon those reported in Emond in that we include all children with pica-reported behaviors in ALSPAC waves through 115 months. This extension improves understanding of which children experience pica behaviors and if those behaviors persist later into childhood. Our study highlights that most pica cases occur outside of autism, as evidenced by the ratio of cases at 36 months: 97% of cases occurred in children without an autism diagnosis (175 cases) compared to 6 cases within autistic children. This demonstrates the clinical significance of our findings, as it suggests that although pica is associated with autism diagnosis, most individuals with pica do not receive an autism diagnosis, indicating the presence of pica across different contexts. Our findings highlight that although pica and autism may be associated, pica can occur outside of autism and developmental disabilities, and that pica is not solely a sub-feature of autism.

In contrast to findings that preschool-aged children with DD did not report elevated pica, current study findings yielded significant associations between DD and pica across all five waves (Fields et al., 2021). This difference is likely explained by differing operational definitions of DD in the present study and the one provided by Fields and colleagues, who used a score of <11 on the Social Communication Questionnaire (SCQ), a measure that is a screening tool for autism. We carefully considered the definition of DD to avoid being overly restrictive and to be consistent with how others defined DD using ALSPAC data (Hameed et al., 2018; Riglin et al., 2017). The prevalence of pica in children with DD reported by Fields et al. is similar to that reported in this study (3.2% vs. 3.6%). The 3.6% prevalence of pica at 36 months observed in our study in children with DD is slightly lower than previous estimates of 4-26% for people with DD (Matson et al., 2013). Again, this is likely explained by the heterogeneity of operational definitions of DD across studies.

Pica behaviors were found to be associated with undereating, overeating, and food fussiness at 36 and 54 months. Identifying these eating behaviors could potentially be a marker of early disordered eating. Previous findings from ALSPAC show that childhood overeating is associated with adolescent binge eating, while undereating and food fussiness in childhood is linked to adolescent anorexia nervosa (Herle et al., 2020). Our results support improved pica screening practices in early childhood (36-54 months) to improve early intervention efforts and potentially mitigate longer-term disordered eating behaviors. One option for screening pica in clinical settings would be the Pica, ARFID, and Rumination Disorder Interview (PARDI; Bryant-Waugh et al., 2019), which is designed for individuals ages 2+ and has demonstrated robust psychometric properties. We did not find evidence that pica behaviors and BMI were associated, similar to findings reported by Hartmann and colleagues (2020), who reported no BMI differences in adults who exhibit long-term pica behaviors relative to those who do not. These findings show that children of all sizes, shapes, and weights may exhibit pica behaviors alongside broader eating difficulties, and this is a salient point for primary care providers who are essential to eating disorder screening and detection (Voss, 2022).

Finally, pica persistence (defined as pica occurrence at two or more waves) was observed in 19.55% of cases. Our findings highlight heterogeneity in pica trajectories. For the majority of children, it is a transient behavior; nonetheless, for a significant subset (19.5%), it persists across development. This is the first study to examine pica recurrence from early childhood (36 months; 3 years) through age 115 months (9 years). It is important to note that individuals must be at least two years old to be diagnosed with pica (Al Nasser, Muco, & Alsaad, 2022). We examined reports of pica behavior beginning at age 36 months, ensuring that early pica behaviors were captured shortly after the required age of diagnosis. More comprehensive longitudinal studies are needed to understand pica behaviors from early childhood into adulthood. A priority of future research is to investigate if those individuals who are reported by their caregivers to exhibit pica behaviors in childhood also report pica behaviors into adolescence and adulthood and whether there are a developmental points where pica is likely to cease and to recur.

Our study has several noteworthy strengths. The data used in this study were obtained from ALSPAC, considered one of the most detailed population-based cohort studies covering childhood to young adulthood (http://www.bristol.ac.uk/alspac/about/). To our knowledge, the present study is the most extensive prospective investigation of pica behaviors spanning a broad timeframe in childhood in a general population sample. This provides greater insight into the longitudinal stability of pica behavior and its association with other co-occurring conditions thereby improving generalizability of findings.

Limitations should also be considered. First, respondent options for indicating pica behavior consisted of “every day,” “at least once a week,” “less than once a week,” and “not at all.” Although this provides insight into the presence of pica behaviors, limits precision of how frequently these behaviors occur within children. For example, a mother who selected “at least once a week” may observe their child eating non-nutritive foods once per week or six times per week. We could not account for the severity of pica behaviors, given the lack of true frequency assessment for pica behaviors. Future studies using population-level data should consider collecting continuous-level data on pica behaviors to better quantify frequency. Dinkler and Bryant-Waugh (2021) noted that pica measurements typically include a single screening item and do not examine specific features of pica. Existing single-item screeners typically inquire into the consumption of non-nutritive foods and contain a Likert-scale, such as that found in the eating Disorders in Youth-Questionnaire (EDY-Q) and Screening Tool of Feeding Problems (STEP) (Matson & Kuhn, 2001; van Dyck & Hilbert, 2016). No screening questionnaires exist that capture all of the diagnostic criteria for pica outlined in the DSM-5. In addition to capturing a more specific frequency of pica behaviors, future work could develop a pica measurement that incorporates multiple items, some of which would permit a better understanding of how pica presents and pica phenotypes. What is observed in this study examining pica in childhood may not translate well into pica observed in later developmental periods and adulthood. There are various causes in adulthood, ranging from co-occurrences with anorexia nervosa, pica triggered by pregnancy, or pica because of factitious disorder (APA, 2013). We observed a decrease in pica behaviors as the child aged. It is unclear if this is because pica prevalence naturally decreased with age or if the caregivers spent less time observing children’s eating behaviors as the children grew older and became more independent. Finally, as this was an exploratory study, we did not correct for multiple comparisons, and some of the observed differences might have arisen by chance.

## Conclusion

This study provides critical information for clinicians to recognize and screen for pica in early childhood regardless of child BMI. Furthermore, providers could screen for pica behaviors in children with autism and DD as pica prevalence was higher (although clearly not limited to) these groups. Ultimately, more tailored and effective diagnostic and treatment options can be developed or adapted through improved understanding of how pica behaviors present and persist throughout the lifespan and its co-occurring conditions.

## Data Availability

The study website contains details of all the available data through a fully searchable data dictionary and variable search tool. To access, reference the following webpage: http://www.bristol.ac.uk/alspac/researchers/our-data/.

## Acknowledgment

“We are extremely grateful to all the families who took part in this study, the midwives for their help in recruiting them, and the whole ALSPAC team, which includes interviewers, computer and laboratory technicians, clerical workers, research scientists, volunteers, managers, receptionists, and nurses.”

